# Adolescents Hospitalized with SARS-CoV-2: Analysis of Clinical Profile and Risk Factors for Severe Disease over a two-year period

**DOI:** 10.1101/2022.01.06.22268674

**Authors:** Tamoghna Ghosh, Tejas M Suri, Kana Ram Jat, Aditya Kumar Gupta, Sushma Bhatnagar, Pawan Tiwari, Saurabh Mittal, Anant Mohan

## Abstract

**Introduction:** There is a lack of studies in adolescents with COVID-19 from developing countries. We aimed to describe the clinical profile and risk factors for severe disease in adolescents hospitalized with COVID-19.

**Methods:** A retrospective analysis of a prospectively admitted cohort of COVID-19 patients was performed at a tertiary hospital in north India. Adolescents aged 12 to 18 years who were hospitalized during the first wave (March 2020 to December 2020) and the second wave (March 2021 to June 2021) of the pandemic were included. Data on the demographic details, clinical presentation, laboratory parameters, disease severity at admission, treatments received, and in-hospital outcomes were retrieved and logistic regression was used to identify the risk factors for occurrence of moderate or severe disease.

**Results:** The study included 197 adolescents with median (IQR) age 15 (13-17) years, of whom 117 (59.4%) were male. Among these, 170 (86.3%) were admitted during the 1^st^ wave. Underlying comorbidities were present in 9 (4.6%) patients. At the time of hospital admission, 60 (30.9%) patients were asymptomatic. In the severity grading, 148 (84.6%) had mild, 16 (9.1%) had moderate, and 11 (6.3%) had severe disease. Fever (14.9%) and cough (14.9%) were the most commonly encountered symptoms. The median (IQR) duration of hospital stay was 10 (8-13) days and 6 (3.1%) patients died in hospital. The odds of moderate to severe disease were 3.8 for second wave, 1.9 for fever and 1.1 for raised C reactive protein (CRP).

**Conclusion:** In our single-center study from northern India, adolescents admitted with COVID-19 had predominantly asymptomatic or mild disease. Admission during the second wave of COVID-19 pandemic, presence of fever and raised CRP were risk factors for moderate or severe disease.

**Lay Summary:** From 3rd January 2022 onwards, adolescents between 15 to 18 years of age in India will be given Covaxin vaccine, as per the latest Indian government guidelines. In our study, we aimed to describe the clinical profile and risk factors for severe disease in adolescents hospitalized with COVID-19. Our study included 197 adolescents. 170 (86.3%) of them were admitted during the 1^st^ wave and the rest 27 (13.7%) during the 2nd wave. At the time of hospital admission, 60 (30.9%) patients were asymptomatic. In the severity grading, 148 (84.6%) had mild, 16 (9.1%) had moderate, and 11 (6.3%) had severe disease. Fever (14.9%) and cough (14.9%) were the most commonly encountered symptoms. The median (IQR) duration of hospital stay was 10 (8-13) days and 6 (3.1%) patients died in hospital. 2nd wave, fever and high C reactive protein increased the odds of moderate to severe disease.

## Introduction

The COVID-19 pandemic has severely affected every aspect of human life and health, including physical, social, behavioural and psychological well-being. Globally, it has affected 288 million people and caused more than 5.4 million deaths as of now (1). Approximately 20% of adult COVID-19 patients require hospitalization, 5% become critically ill, and between 2 to 5% may die (1). Although children constitute a relatively small proportion of the hospitalized COVID-19 cases with relatively lower severity and mortality, there has been a surge in the number of cases in children with the progression of the pandemic (2).

Various vaccines have been developed to counteract COVID-19, but till now the vaccines available in India were restricted for use among adults (aged 18 years and above) only. However, from 3rd January 2022 onwards, adolescents between 15 to 18 years of age will be given Covaxin vaccine, as per the latest government guidelines (3). India has the largest adolescent population in the world of approximately 253 million, and every fifth person is between 10 to 19 years. Adolescents in the age group of 12 to 18 years have already suffered due to closures of schools during the lockdowns hampering their educational, emotional and social development (4). Worryingly, reports suggest that adolescents are also susceptible to developing severe COVID-19 illness. (5).

Although there have been multiple studies involving adults and children below 12 years of age (6), the clinical characterization and course of covid-19 in the 12-18 age adolescent group is still not well defined.

Further, there are limited studies assessing risk factors for severe disease, particularly from developing countries. This study aimed to describe the clinical profile and risk factors for severe disease in adolescents hospitalized with COVID-19 in a tertiary care teaching hospital from India.

## Materials and Methods

### Study population

We performed a retrospective analysis on a prospectively enrolled cohort of patients admitted at a dedicated COVID-19 tertiary care hospital in North India. We retrieved the clinical data of adolescents aged 12 to 18 years hospitalized with SARS-CoV-2 during the first (March 2020 to December 2020) and second (March 2021 to June 2021) waves of COVID-19 pandemic in India. Patients with a positive RT-PCR or cartridge-based nucleic acid amplification test (CBNAAT) for SARS-CoV-2 from secretions of the upper or lower respiratory tract were eligible for inclusion in the study. Tests for SARS-CoV-2 were performed as per the Indian Council of Medical Research (ICMR) guidelines (7). Besides, all hospitalized patients were tested for SARS-CoV-2 as per institutional policy. Initially, all cases of COVID-19 were being hospitalized, however, from July 2020,the Government of India recommended home isolation for patients who were asymptomatic or mildly symptomatic without any comorbidity. Consequently, only moderate to severe cases, adolescents with comorbidities and those for whom home isolation was not feasible were hospitalized. We excluded home isolated patients from our study.

### Data collection

We extracted data from the electronic records of patients regarding demographic details; clinical features including nature, duration and severity of symptoms; various laboratory parameters; and patients’s in-hospital outcomes (discharge or death). The patients were treated according to the institutional protocols. In most cases, treatment was supportive, and specific medications were used on a case-to-case basis. Outcome measures included descriptive statistics related to demographic, clinical and laboratory parameters in the patients. We classified disease severity as per WHO guidelines: patients were deemed to have severe disease if SpO_2_ was less than 90% or respiratory rate was greater than 30 per minute; moderate disease if there were symptoms of pneumonia (fever, cough, tachypnea and breathlessness) with SpO_2_ greater than 90%; and mild disease if there were no symptoms or signs of pneumonia or hypoxemia (9). The study protocol was approved by the institute ethics committee (IEC).

### Statistical analysis

Data was analysed using Stata v14 (StataCorp LP, College Station, TX). Categorical data was reported as a percentage (%). Continuous parameters were reported as mean (standard deviation, SD) if data were normally distributed, or median (interquartile range, IQR) if data had a skewed distribution. The patients were classified into two groups to assess risk factors for disease severity: the first group included asymptomatic and mild illness; and the second group included moderate, and severe illness. Clinical and laboratory parameters were compared among both groups using bivariate analysis. Means were compared using Student’s t-test and medians using Mann–Whitney test. Logistic regression was performed to identify risk factors for moderate or severe disease. Factors which had p-value less than 0.1 in bivariate analysis were considered for logistic regression. p-value of less than 0.05 was considered as significant.

## Results

### Demography

We enrolled a total of 197 adolescents aged 12 to 18 years in the study. Demographic details of included patients are shown in Table 1. The median (IQR) age was 15 (13-17) years with 117 (59.4%) males. Among these, 170 (86.3%) were enrolled during the first wave and 27 (13.7%) were enrolled during the second wave of the COVID-19 pandemic in India. A total of 9 (4.6%) patients had underlying comorbidities, with solid organ malignancy, rheumatic disorder and hypothyroidism being the most common.

**Table 1:** Demographic and clinical characteristics of adolescents with SARS-CoV-2.

| Characteristics (data available) | Value |
| --- | --- |
| Age (years), median (IQR) (n=197) | 15 (13-17) |
| Gender, male:female (n=197) | 117:80 |
| Wave, first:second (n=197) | 170:27 |
| Comorbidities |  |
| Diabetes mellitus, n (%) (n=197) | 2 (1.0) |
| Hypertension, n (%) (n=197) | 1 (0.5) |
| Coronary Artery Disease, n (%) (n=197) | 2 (1.0) |
| Hypothyroidism, n (%) (n=197) | 3 (1.5) |
| Asthma, n (%) (n=197) | 2 (1.0) |
| Chronic Kidney Disease, n (%) (n=45) | 2 (4.4) |
| Rheumatologic disorder, n (%) (n=42) | 4 (9.5) |
| Hematological malignancy, n (%) (n=41) | 2 (4.9) |
| Solid organ malignancy, n (%) (n=47) | 6 (12.8) |
| Duration of symptoms, days, median (IQR) (n=132) | 3 (2-5) |
| Symptoms |  |
| Asymptomatic at admission, n (%) (n=194) | 60 (30.9) |
| Fever, n (%) (n=194) | 29 (14.9) |
| Nasal symptoms, n (%) (n=194) | 17 (8.8) |
| Cough, n (%) (n=194) | 29 (14.9) |
| Sputum, n (%) (n=194) | 4 (2.1) |
| Dyspnoea, n (%) (n=194) | 12 (6.2) |
| Fatigue, n (%) (n=192) | 20 (10.4) |
| Myalgia, n (%) (n=192) | 22 (11.5) |
| Diarrhoea, n (%) (n=192) | 5 (2.6) |
| Nausea/vomiting, n (%) (n=192) | 3 (1.6) |

IQR, interquartile range

### Clinical features

Among the enrolled patients, 60 (30.9%) were asymptomatic at hospital admission. In the severity grading, 148 (84.6%) had mild, 16 (9.1%) had moderate, and 11 (6.3%) had severe disease. Fever and cough were the most commonly encountered symptoms, and were present in 14.9% patients each. Other common symptoms were myalgia, fatigue, nasal symptoms and dyspnoea (Table 1). The median (IQR) duration of symptoms at presentation was 3 (2-5) days. Table 2 summarizes the laboratory investigations in included patients with the proportion of patients having abnormal values. Common laboratory abnormalities included high ferritin (34.9% of patients), high D-dimer (15.5%), low alkaline phosphatase(57.8%), low platelet count (19.3%), low total leukocyte count (21.1%), high NLR (15.1%), and low hemoglobin were the common laboratory abnormalities.

**Table 2:** Laboratory parameters in adolescents infected with SARS-CoV-2.

| **Laboratory parameter**<br>**(data available)** | **Median**<br>**(IQR)/mean**<br>**(SD)** | **Abnormal**<br>**Values** | **Proportion of children**<br>**having abnormal values,**<br>**n (%)** |
| --- | --- | --- | --- |
| Haemoglobin (Hb), g/dl<br>(n=176) | 12.3 (2.4) | Hb <11.5 | 53 (30.1) |
|  |  | Hb <7.0 | 8 (4.6) |
| Total leucocyte counts<br>(TLC)/mm<sup>3</sup> (n=175) | 5400 (4070-6730) | TLC <4000 | 37 (21.1) |
|  |  | TLC >12000 | 4 (2.3) |
| Neutrophil, % (n=173) | 48 (42-59) | Neutrophils<br><40% | 34 (19.7) |
| Lymphocytes, %<br>(n=172) | 37 (29-43) | Lymphocytes<br><25% | 35 (20.4) |
| NLR (n=172) | 1.3 (1.0-2.1) | NLR> 3 | 26 (15.1) |
| Platelet counts/mm<sup>3</sup><br>(n=176) | 234000 (171500-300000) | Platelets <150<br>000 | 34 (19.3) |
| Urea, mg/dl (n=176) | 19.3 (14.2-23.5) | Urea >40 | 9 (5.1) |
| Creatinine, mg/dl<br>(n=176) | 0.58 (0.45-0.68) | Creatinine >0.9 | 6 (3.4) |
| Total bilirubin, mg/dl<br>(n=176) | 0.6 (0.4-0.8) | Bilirubin >1 | 25 (14.2) |
| SGOT, IU/dl (n=173) | 29 (23-37) | SGOT >45 | 26 (15.0) |
| SGPT, IU/dl (n=173) | 22 (15-35) | SGPT >45 | 29 (16.8) |
| Alkaline phosphatase<br>(ALP), IU/dl (n=173) | 120 (90-189) | ALP >140 | 73 (42.2) |
| Total protein, g/dl<br>(n=174) | 6.9 (0.7) | Total protein <6.1 | 11 (6.3) |
| Serum albumin, g/dl<br>(n=174) | 4.4 (0.5) | Albumin <3.5 | 9 (5.2) |
| Ferritin, ng/ml (n=89) | 37 (21-107) | Ferritin >60 | 31 (34.8) |
| C-reactive protein<br>(CRP), mg/L (n=121) | 0.11 (0.01-0.73) | CRP >6 | 11 (9.1) |
| Fibrinogen, mg/dl<br>(n=108) | 285 (249-329) | Fibrinogen>400 | 10 (9.3) |
| D-dimer, ng/ml (n=116) | 107 (69-250) | D-dimer >500 | 18 (15.5) |

SARS-CoV-2, severe acute respiratory syndrome coronavirus-2; IQR, interquartile range;
SD, standard deviation; NLR, neutrophil lymphocyte ratio; SGOT, serum glutamic
oxaloacetic transaminase; SGPT, serum glutamic pyruvic transaminase

### Treatment and in-hospital outcomes

The majority of symptomatic patients received supportive therapy. Asymptomatic patients did not receive any treatment. Some patients received additional therapies, as shown in Table 3. The most commonly used drugs were oral vitamin C, antipyretics, and antihistamines. Fourteen patients received steroids and 13 patients received oxygen. The median (IQR) duration of hospital stay was 10 (8-13) days. The majority of the patients survived; 6 (3.1%) patients died.

**Table 3:** Treatment received in adolescents infected with SARS-CoV-2.

| **Nature of treatment (data available)** | **Number (%) of children, received the treatment** |
| --- | --- |
| Antipyretics (n=177) | 126 (71.2) |
| Antihistamines (n=177) | 112 (63.3) |
| Oral Vitamin C (n=177) | 173 (97.9) |
| Teicoplanin (n=175) | 7 (4.0) |
| Dalteparin (n=175) | 18 (10.3) |
| Remdesivir (n=59) | 10 (16.9) |
| Steroid (n=58) | 14 (24.1) |
| Tocilizumab (n=177) | 1 (0.6) |
| Oxygen (n=177) | 13 (7.3) |
| NIV/HFNC (n=177) | 5 (2.8) |
| Intubation (n=171) | 4 (2.3) |

SARS-CoV-2, severe acute respiratory syndrome coronavirus-2; NIV, non-invasive ventilation; HFNC, high-flow nasal cannula

### Risk factors for moderate-severe disease

On bivariate analysis, the risk factors for moderate or severe disease included lower age, admission during India’s second wave of COVID-19 pandemic, solid organ malignancy, greater symptom duration at admission, lower hemoglobin level, lower serum albumin level, higher C reactive protein (CRP) level, higher D dimer level, and steroid or oxygen administration (Table 4). A total of 5 (18.5%) patients with moderate-severe disease died, whereas 1 (0.7%) patient with mild disease died. The results of logistic regression are shown in Table 5. On regression analysis, the significant risk factors for moderate or severe disease included admission during the second wave of COVID-19 pandemic, presence of fever, and high CRP level (greater than 6 mg/L).

**Table 4:**
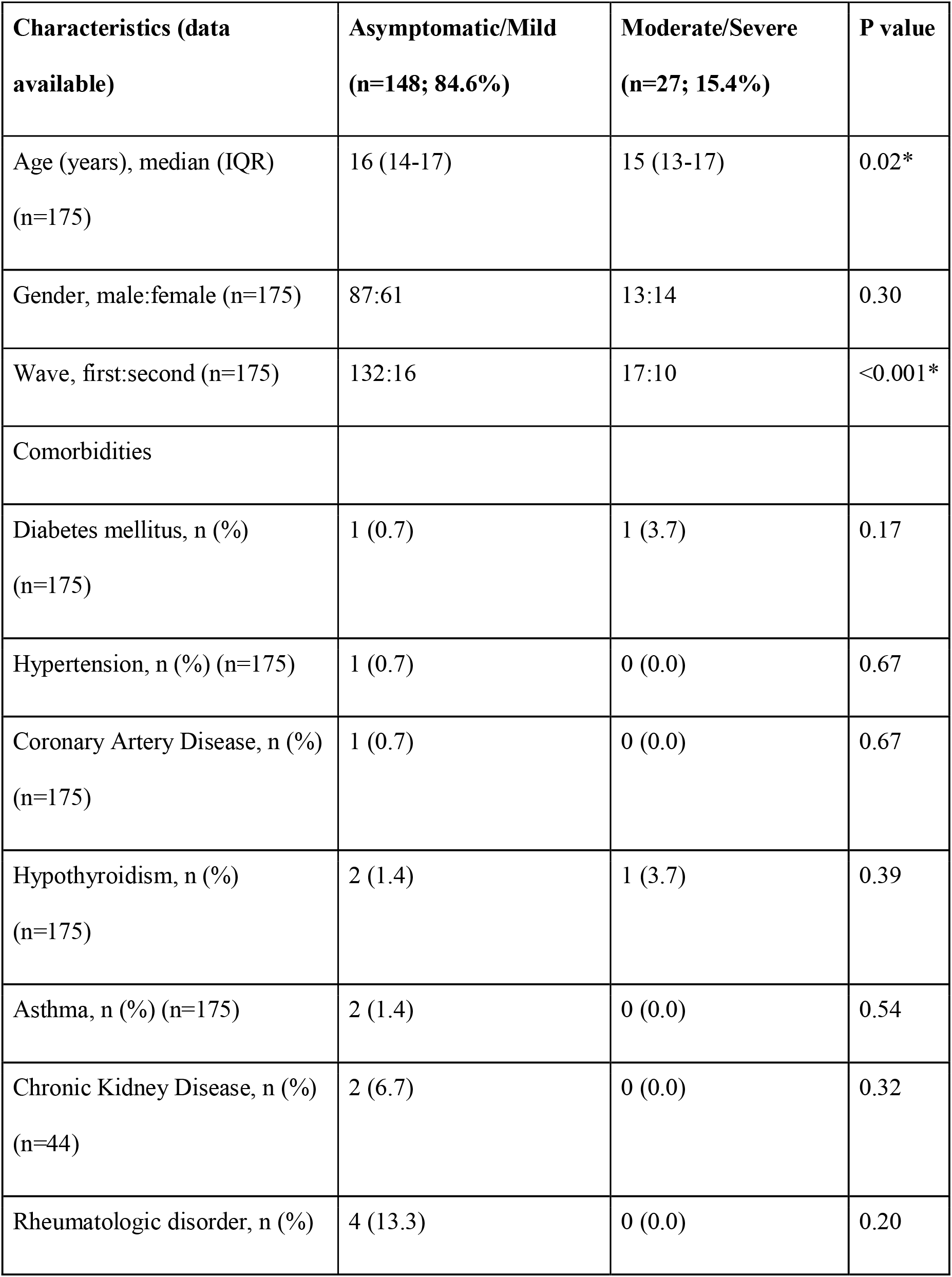
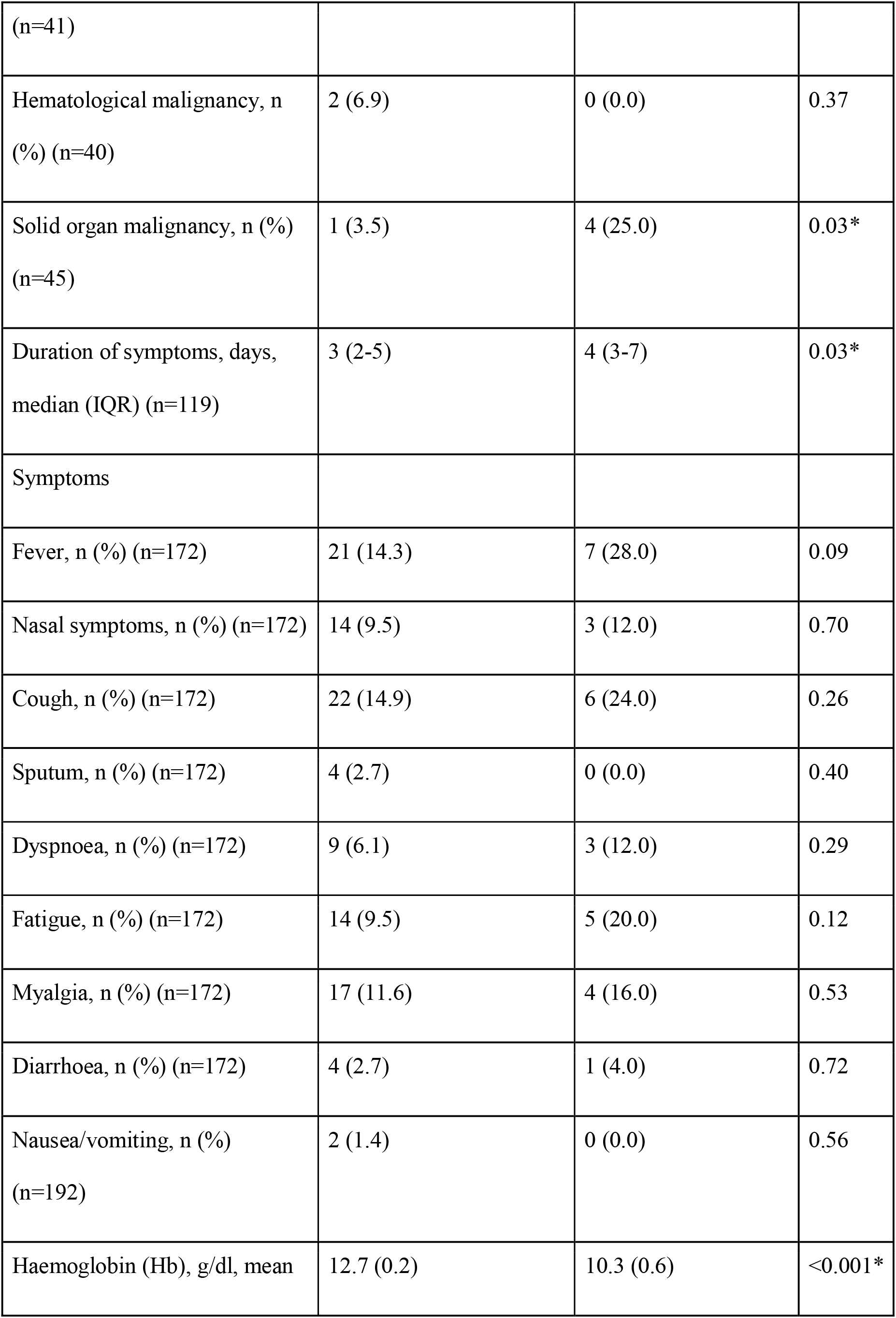
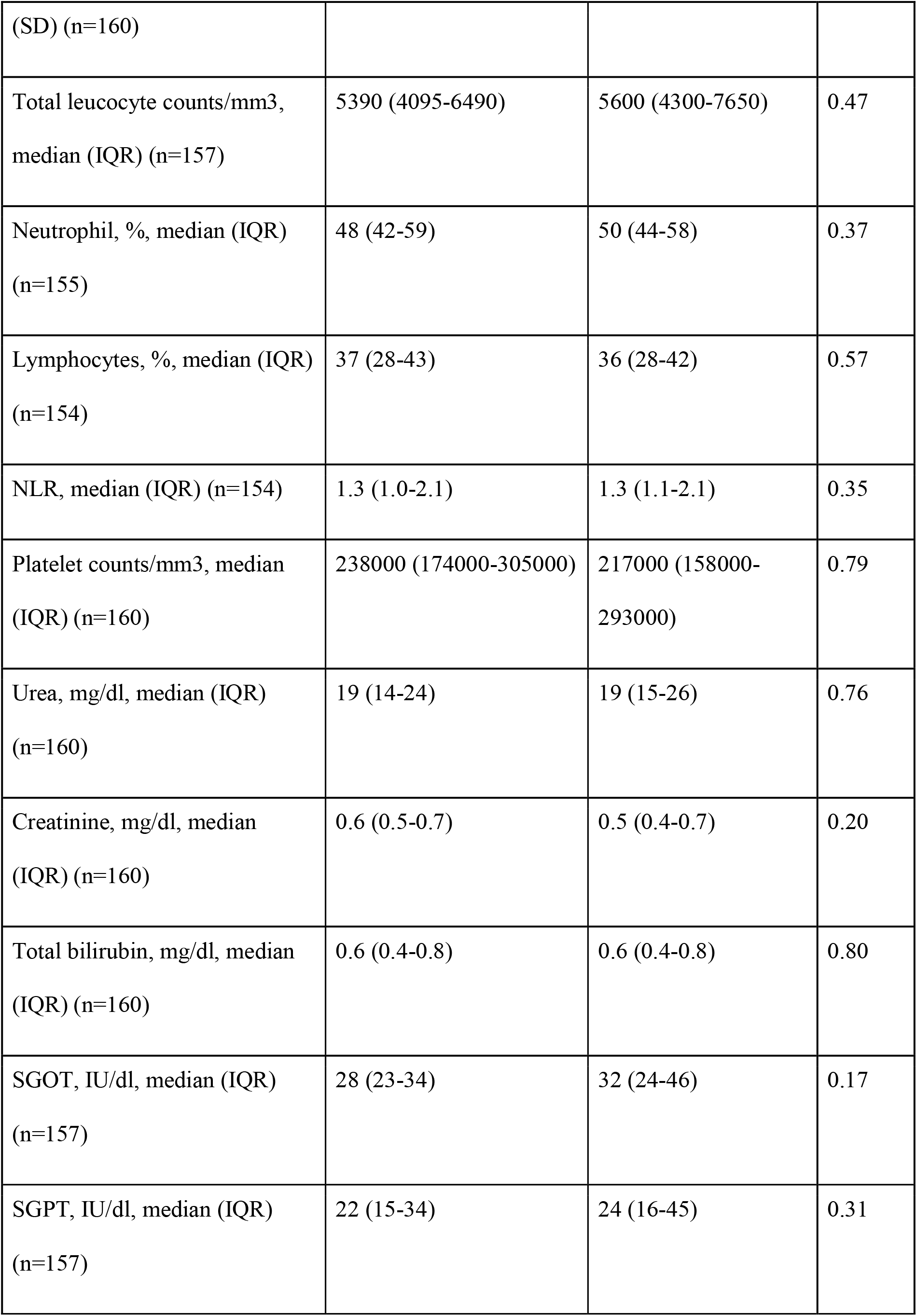
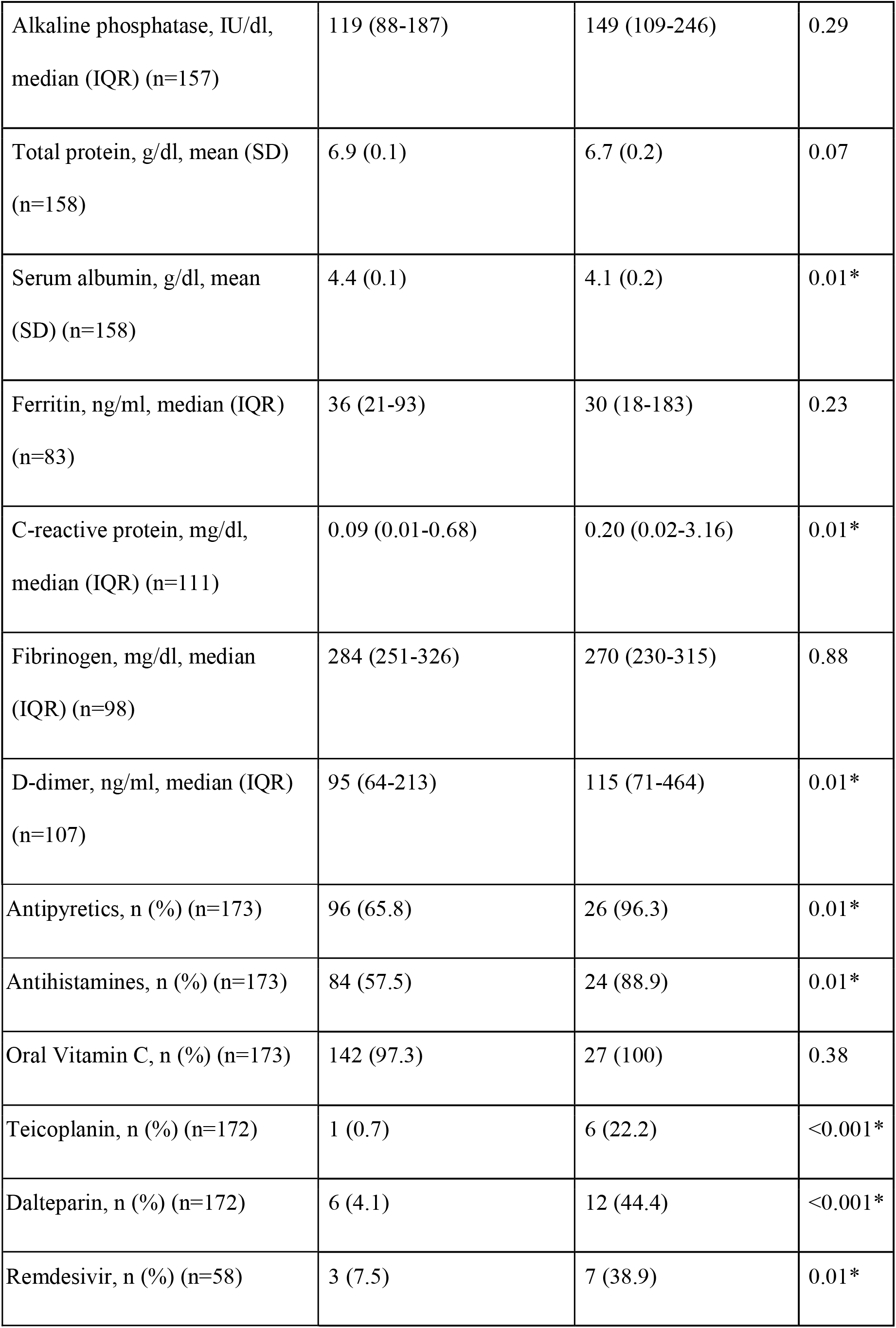
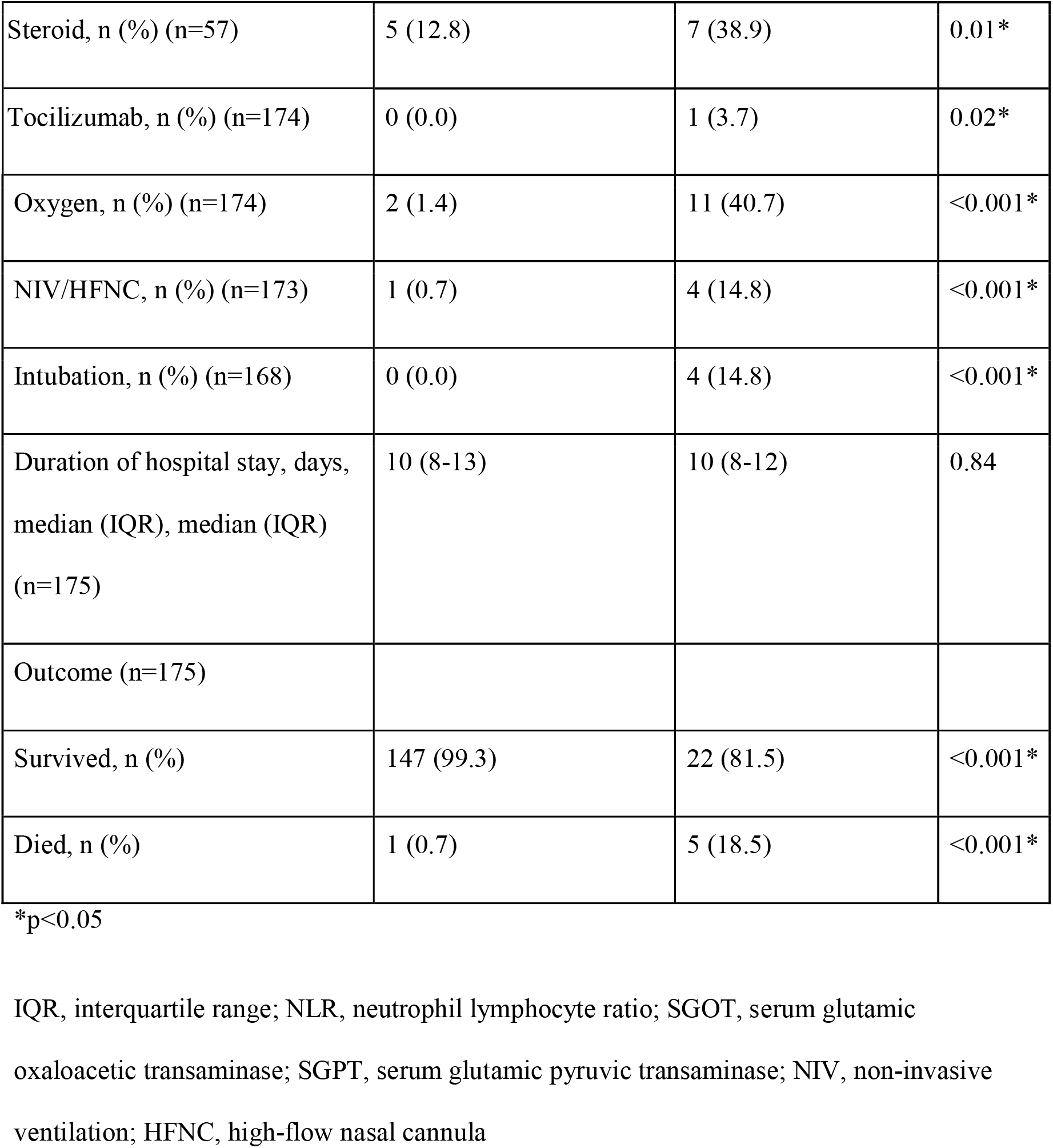
Clinical features for Asymptomatic/Mild vs Moderate/Severe cases of COVID-19 in adolescents.

**Table 5:** Logistic regression of features associated with moderate-severe disease in adolescents with COVID-19.

| **Risk factor** | **Coefficient (95% CI)** | **Odds Ratio (95% CI)** | **p value** |
| --- | --- | --- | --- |
| Age | -0.37 (-0.89- 0.15) | 0.69 (0.41- 1.17) | 0.16 |
| 2nd Wave | 1.35 (1.11- 3.80) | 3.84 (1.33- 44.76) | 0.03* |
| Fever | 0.62 (0.16- 3.03) | 1.86 (1.17- 20.67) | 0.01* |
| Hemoglobin | -0.20 (-0.55- 0.16) | 0.82 (0.57- 1.17) | 0.27 |
| Serum albumin | 0.13 (-2.72- 2.98) | 1.14 (0.07- 19.78) | 0.93 |
| D dimer | 0.00 (0.00- 0.01) | 1.00 (0.99- 1.01) | 0.81 |
| C Reactive Protein | 0.08 (0.02- 0.40) | 1.09 (1.02- 1.49) | 0.04* |
| Total Protein | 0.87 (-1.30- 3.04) | 2.38 (0.27- 20.88) | 0.43 |

\*p<0.05
CI, confidence interval

## Discussion

In this retrospective analysis of a prospectively enrolled cohort from a dedicated COVID-19 hospital in North India, we described the clinical and laboratory profile of 197 adolescents aged 12 to 18 years hospitalized with COVID-19 infection. A large proportion of the subjects were asymptomatic or had mild disease at hospital admission. Admission during the second wave of COVID-19 pandemic in India, presence of fever, and high CRP value were significant risk factors for moderate or severe disease.

Fever and cough were the most common encountered symptoms in our patients. Previously, a multicentre study of adolescents with COVID-19 in Italy reported fever (82.1 %) to be the most common symptom (13). Similarly, a systematic review of studies enrolling children with COVID-19 found fever and cough to be the most common symptoms (14). In contrast, other studies conducted in Chinese and American children with COVID-19 found fever to be less common (36–56%) compared with cough or pharyngitis (10–12). Although there are no previous Indian studies describing clinical features of COVID-19 among adolescents, fever and cough have been reported to be most common symptoms of Indian COVID-19 patients in both the under-12 years age group (6) and in adults (15).

In a previous Indian multicentre study conducted among children under the age of 12 years hospitalized with COVID-19, the most common laboratory abnormalities included lymphopenia, thrombocytopenia, low alkaline phosphatase and high ferritin (6). We observed similar findings in adolescents aged 12 to 18 years. Additionally, we found raised D-dimer levels in a substantial proportion. Although the effects of SARS-CoV on hematopoiesis are still being explored, it has been proposed that the virus-mediated infection leads to consumption of T-lymphocytes, particularly the CD4 and CD8 T-cells, thereby leading to lymphopenia (16). For thrombocytopenia, a proposed mechanism is virus-mediated endothelial damage leading to platelet activation and micro thrombus formation in the pulmonary vasculature, which in turn leads to platelet consumption (17). Meanwhile, higher serum ferritin levels have been found to be associated with severe pulmonary involvement, independent of age and gender (18). D-dimer is an indicator of activation of the coagulation cascade and the fibrinolytic system. This activation of the coagulation cascade is postulated to be due to the viremia, accentuated cytokine levels, infection, and organ dysfunction (19).

We observed a relatively low in-hospital mortality rate (3.1%) in our cohort. This is in concurrence with other studies which observed low mortality in younger age groups (2) with mortality gradually increasing with age (20). This can be attributed to lower comorbidity, better immune response, and lesser exposure in adolescents in comparison to adults and geriatric population (21).

In our study, admission during the second wave of the pandemic in India was a significant risk factor for severe disease and increased mortality. During this wave, there was the emergence of the SARS-CoV-2 B.1.617.2 (delta) variant, which was more transmissible, more likely to evade humoral immunity (to vaccination or previous infection), and more likely to cause severe disease in comparison to the previous strain of the virus seen in the first wave (22).

The strength of this study is that it is one of the largest single centre studies on COVID-19 in the adolescent age group in India, covering both the waves of the pandemic. Secondly, a comprehensive clinical and laboratory spectrum was recorded and analyzed. Inflammatory markers in the adolescent age group had been rarely reported. However, this study was limited due to its retrospective design. Further, laboratory tests were performed mostly in patients with greater severity of illness which may lead to overestimation of laboratory abnormalities in this study compared to community-based studies or studies enrolling both hospitalized and non-hospitalized patients of similar demography.

In conclusion, we found that among adolescents admitted due to COVID-, the majority had asymptomatic and mild illness. The presence of fever, high CRP level and admission during the second wave of the pandemic were risk factors for moderate or severe illness.

## Data Availability

Data will be made available on reasonable request.

## Acknowledgements

The authors express their sincere gratitude to the Data Entry Staff of Pulmonary, Critical Care, and Sleep Medicine, All India Institute of Medical Sciences, New Delhi, India, for their assistance in procuring the clinical and laboratory details and preparing the raw data.

## Availability of data and materials

The data would be made available by the authors on specific request keeping patient confidentiality in view.

## Ethics approval

The study was approved by the Institute Ethics Committee, AIIMS, New Delhi, India.

## Authors’ contributions

All listed authors meet the ICMJE criteria. We attest that all authors contributed significantly to the creation of this manuscript, each having fulfilled criteria as established by the ICMJE.

